# Ten years of population-level genomic *Escherichia coli* and *Klebsiella pneumoniae* serotype surveillance informs vaccine development for invasive infections

**DOI:** 10.1101/2020.07.08.20140707

**Authors:** Samuel Lipworth, Karina-Doris Vihta, Kevin K Chau, James Kavanagh, Timothy Davies, Sophie George, Leanne Barker, Ali Vaughan, Monique Andersson, Katie Jeffery, Sarah Oakley, Marcus Morgan, Timothy EA Peto, Derrick W Crook, A Sarah Walker, Nicole Stoesser

## Abstract

The incidence of bloodstream infections (BSIs) caused by *Enterobacteriaceae* (e.g. *Escherichia coli, Klebsiella pneumoniae*) continues to increase globally and the threat of untreatable disease is substantial^1^. Prophylactic vaccines represent an alternative approach to combating antimicrobial resistance (AMR) by reducing antibiotic usage and preventing infections caused by AMR-associated strains. To investigate their potential utility, we performed *in silico* serotyping on 4035 *E. coli*/*K. pneumoniae* BSI from population-level surveillance in Oxfordshire (2008-2018) in addition to 3678 isolates from previous studies. Most infections, including those associated with AMR, were caused by isolates with a small subset of O-antigens, with no evidence that the proportion of BSIs caused by these changed significantly over time. O-antigen targeted vaccines might therefore be useful in reducing the significant morbidity and mortality^2^ associated with BSIs. Vaccines may also have a role in preventing the spread of carbapenem resistance genes into common serotypes associated with community-onset disease.

---

For *E. coli*, the O-antigen, a component of lipopolysaccharide (LPS), has been considered the most promising vaccine target^3^. A recently developed bioconjugate vaccine (ExPEC4V; Janssen Pharmaceuticals) targeting four *E. coli* O-antigens (O1A, O2, O6A and O25B) has subsequently shown promise in a Phase II study^4^. For *K. pneumoniae*, vaccine development is lagging behind^5^, and there are no vaccines currently in clinical trials. Large scale, systematic typing studies could help to inform the rational selection of vaccine targets across population groups which best mitigate the risk of BSI (including those associated with AMR) and represent a baseline for monitoring the emergence of serotype variation in response to vaccine roll-out. Most existing studies have been either small, selective, historic (i.e. earlier than 2010), or over limited timeframes in any given setting.

To assess the diversity of antigens, and therefore how effective a vaccine targeted against these might be in reducing the incidence of invasive disease, we sequenced all (90-day patient-deduplicated) isolates from patients presenting with *E. coli* or *K. pneumoniae* (including the closely related *quasipneumoniae* subspecies) BSIs to Oxford University Hospitals between 2008-2018. *In silico* serotyping was performed using the Ectyper tool^6–8^ for *E. coli* and with Kleborate for *Klebsiella spp*.^*9*^ Data was available through the Infections in Oxfordshire Research Database.

For comparisons with existing studies, *in silico* antibiotic resistance gene detection (as called by Kleborate) was used instead of *in vitro* phenotyping because the latter were not universally available. Stacked negative binomial regression was used to compare incidence rate ratios over time (per year longer, IRRy) of BSI belonging to different antigen groups^10^. Healthcare-associated (HA) infection was defined as either nosocomial infection (onset >48 hours after admission) or onset within 30 days of last discharge and community-associated (CA) as that occurring >30 days since last admission.

A total of 3479 *E. coli* isolates were sequenced of which 3278 passed *in silico* serotyping quality control. From these, 106 unique O-antigens were identified, though a relatively small number of these accounted for the majority of isolates (Fig.1). The four most common serotypes (O1A, O2, O6A and O25B; i.e. those covered by the ExPEC4V vaccine) were identified in 1499/3278 (46%) isolates. Their incidence increased over time (IRRy=1.17; 95% CI:1.10-1.24); as did non-ExPEC4V serotypes (IRRy=1.17; 95% CI:1.08-1.27) (p_heterogeneity_=0.6). There was no evidence that O1/O2/O6/O25 serotypes differed in prevalence between CA and HA isolates (912/1969, 46% vs 576/1292, 45% respectively; p=0.3), or across five age-groups (<18 years 52/122 (43%), 18-39 years 98/222(44%), 40-59 years 204/477 (43%), 60-79 years 577/1282 (45%), >80 years 557/1158(48%); p=0.3).

**Figure 1:**
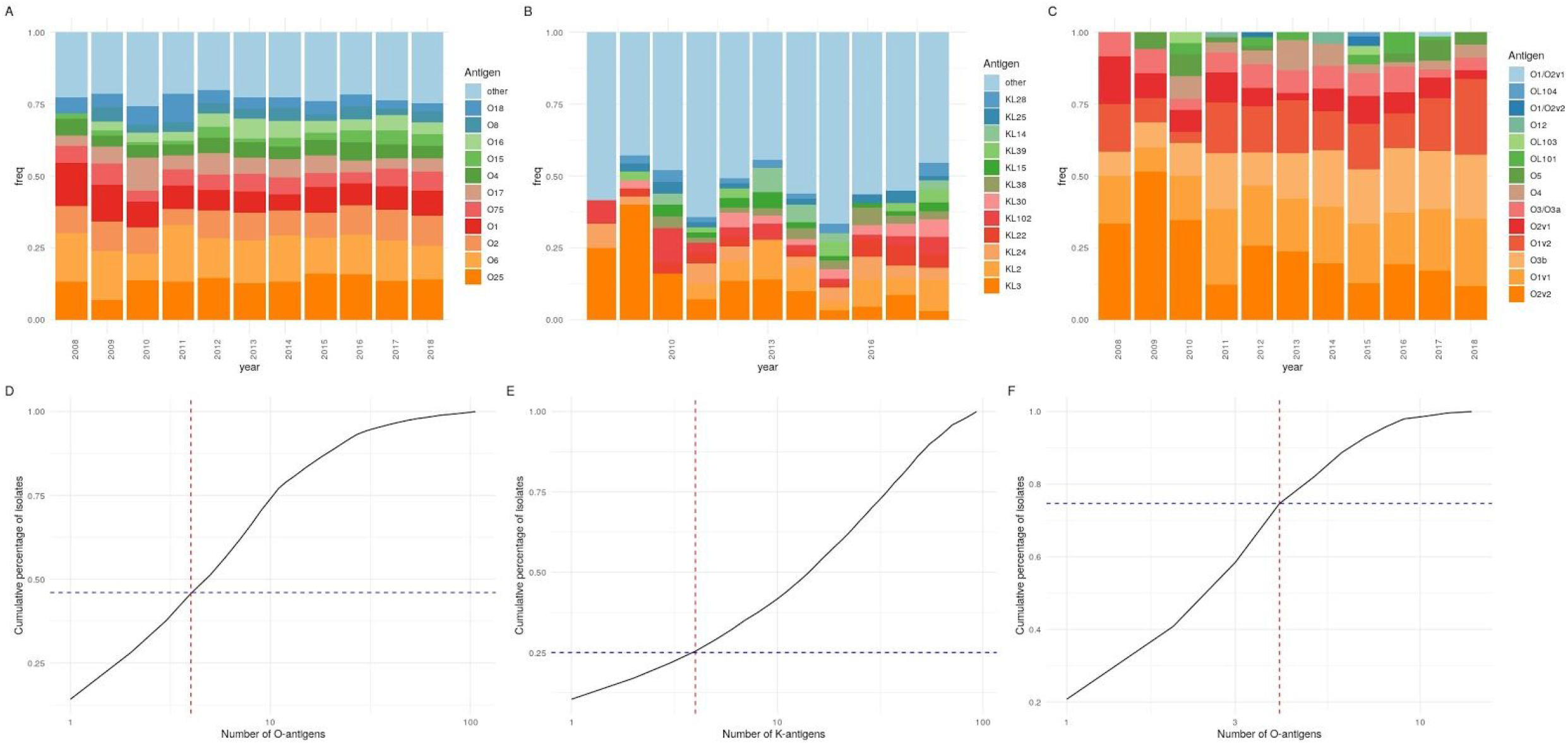
Limited O-antigen diversity in Oxfordshire BSIs with broadly stable population structure over time. Proportion of A. *E. coli* O-antigens B. *Klebsiella* spp. capsular antigens and C. *Klebsiella* spp. O-antigens observed in all isolates over the ten year period. In D.-F., The red dashed line denotes the x-axis position of the four most prevalent O/K-types and the blue dashed line demonstrates the cumulative percentage of isolates which have these antigens, for *E. coli* O-antigens (D.), *Klebsiella* spp. capsular (K) antigens (E.), and *Klebsiella* spp. O-antigens (F.)

Of the 3251 isolates for which in vitro antimicrobial sensitivity data was available, the ExPEC4V serotypes accounted for 191/331 (58%) of extended-spectrum beta-lactamase (ESBL)-producing and 394/834 (47%) of multi-drug resistant (MDR; resistance to ≥3 antibiotic classes) infections. O25 was particularly associated with both ESBL (162/450, 36%) and MDR (249/450, 55%) phenotypes (Fig.2).

**Figure 2:**
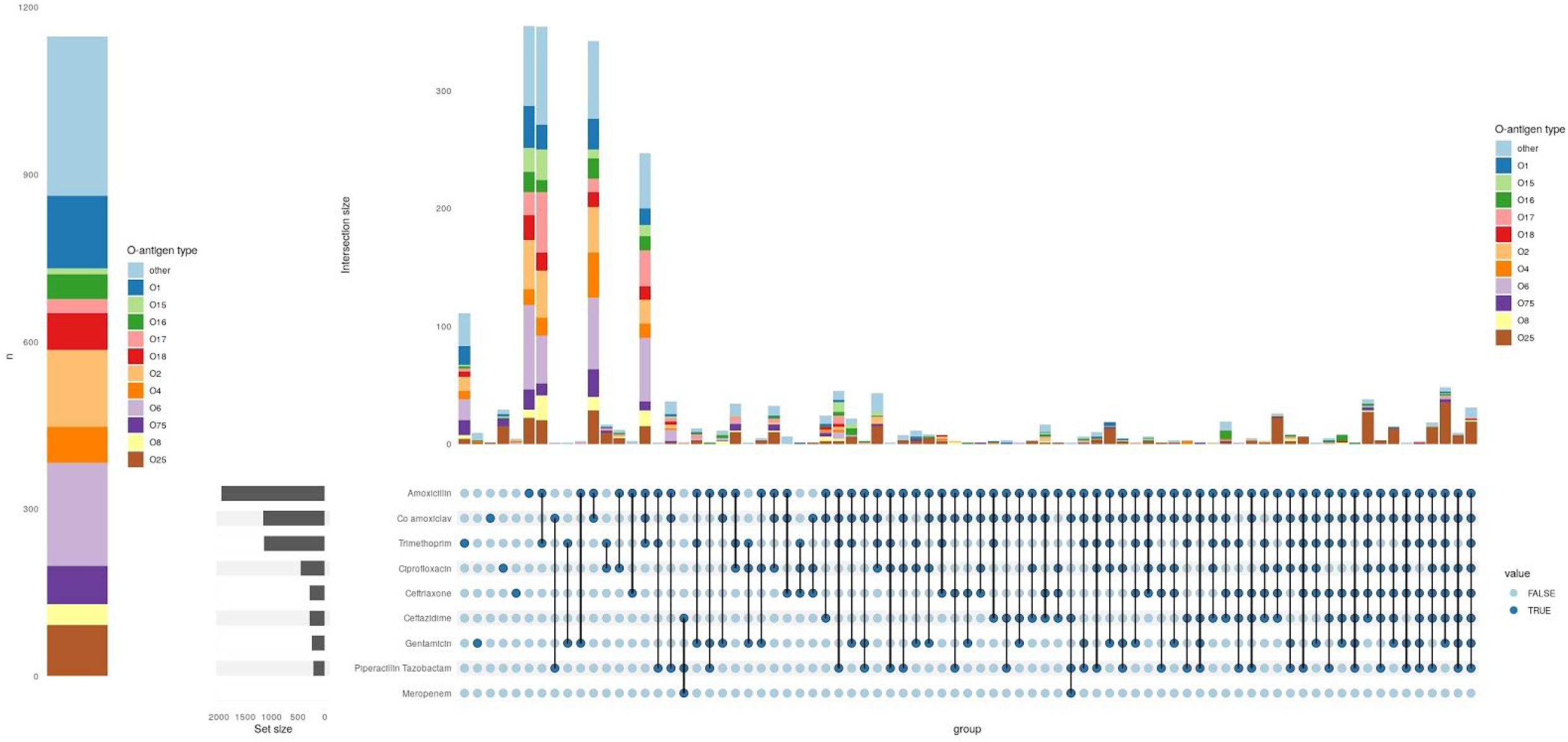
Left: number of fully-sensitive (to antibiotics shown in the right-hand upset plot) *E. coli* BSI isolates by O-type, Right: Upset plot showing the number of *E. coli* isolates with phenotypic resistance to the antibiotics shown, stratified by O-antigen type (colours) and ordered by number of intersections. The horizontal bar plot shows the total number of isolates phenotypically resistance to each antibiotic in any combination.

Similarly we sequenced 556 *K. pneumoniae (and quasipneumoniae)* isolates; 93 unique K- and 14 unique O-loci were confidently identified in 535 (K-antigen)/549 (O-antigen) isolates passing quality control. To determine the theoretical feasibility of developing a *K. pneumoniae* vaccine and for comparison with *E. coli*, we again considered the top four K- and O-loci. KL2, KL22, KL24 and KL3 were identified in 136/535 (25%) isolates. There was no evidence that the incidence of BSIs caused by this group of capsular types changed over time (IRRy=1.05; 95%CI:0.94-1.17), nor that incidence trends varied between these and other *Klebsiella* spp. BSIs (p_heterogeneity_=0.15). By contrast, O2v2, O1v1, O3b and O1v2 accounted for 410/549 (75%) isolates and the incidence of BSIs caused by these increased over time (IRRy=1.12; 95%CI:1.04-1.21), though again there was no evidence that these incidence trends differed from all other O-types (p_heterogeneity_=0.5).

KL3 and KL30 were the most common capsular antigens associated with the ESBL phenotype (43/123 [35%] and 12/123 [10%] respectively). KL3 was also the K-antigen most commonly associated with MDR (45/148 [30%]). In total the four most common K-antigens overall (KL2/242/24/3) accounted for 50/123 (41%) of ESBL and 65/148 (44%) of MDR isolates. In contrast the four most common O-antigens accounted for 106/123 (86%) of ESBL and 122/148 (82%) MDR isolates, with no evidence that the proportion of HA/CA BSI caused by these four antigens differed (179/238 [75%] vs 230/310 [74%], respectively; p=0.9).

Previous studies have suggested that clinically hypervirulent *K. pneumoniae* isolates (i.e. causing CA invasive infection) are particularly associated with the KL1/2 capsular types^11^. There were only two isolates with the KL1 capsular antigen in our study, only one of which carried the virulence-associated genes encoding for yersiniabactin, colibactin, aerobactin, salmochelin, and *rmpA*. Whilst KL2 did have the most isolates carrying hypervirulence genes of any capsular type, there were also 13/35 KL2 isolates with no virulence loci (Fig.3). 36/37 KL1/2 isolates had O1v1 or O1v2 antigens (the other had O1/O2v2) meaning that a quadrivalent O-antigen-targeted vaccine would theoretically provide good coverage against this important group of isolates. Whilst there were more CA than HA isolates for the KL2 capsular type (19/15), this was not the only K-type where this was true and the clinically hypervirulent phenotype was observed in 74 capsular types in total.

**Figure 3:**
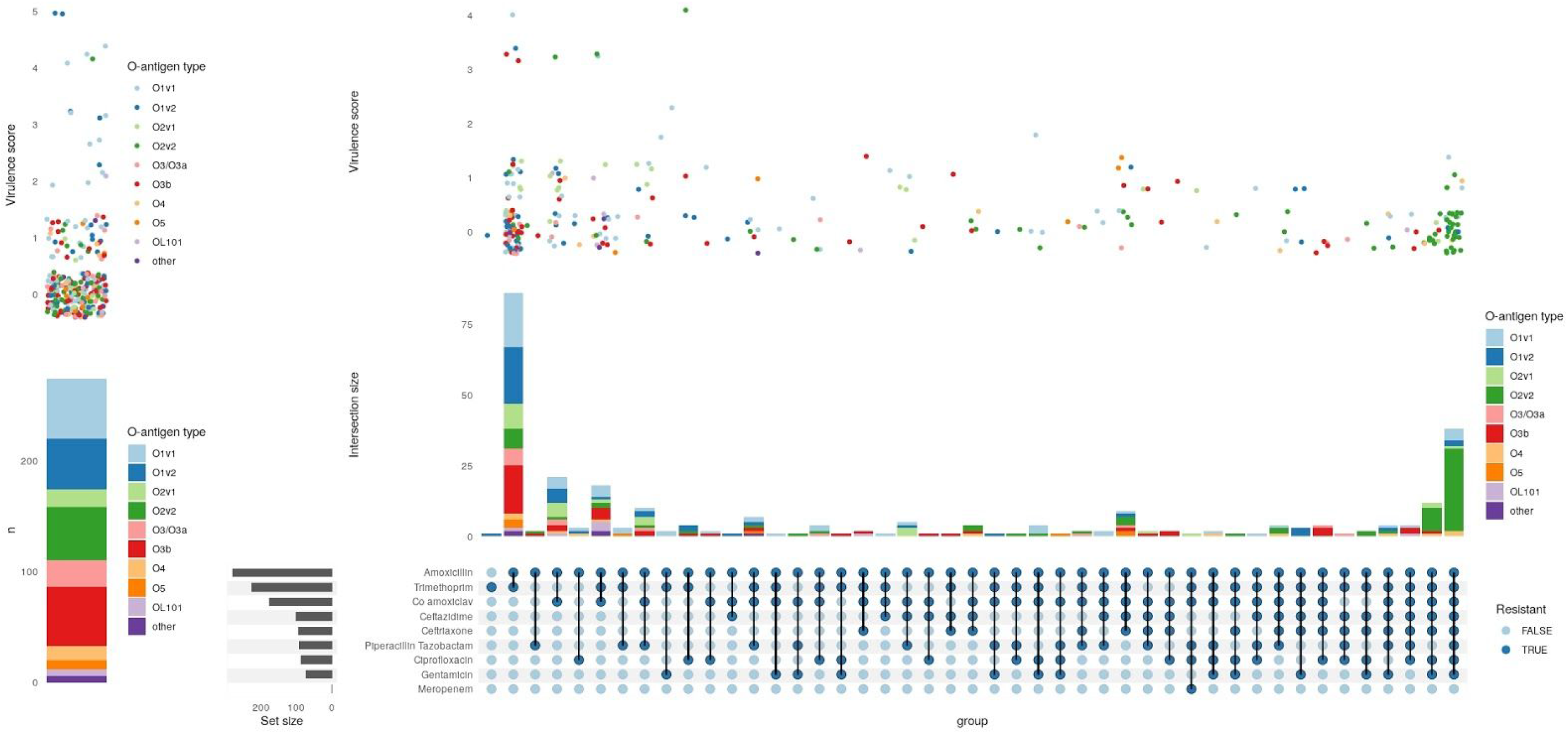
Left: number of *K. pneumoniae* BSI isolates either fully susceptible or resistant only to amoxicillin shown by O-antigen type. The above violin/jitter plot above shows the distribution of virulence scores, points are coloured according to their O-antigen type. Right: Upset plot showing the number of *K. pneumoniae* isolates with resistance to the classes of antibiotics shown. The stacked bar plot shows the distribution of these by O-antigen type and the violin plot (with jittered points to show individual data points) above this demonstrates the corresponding distributions of virulence scores.

As the isolates from our study originated from a single region of the UK with a relatively low AMR burden, we compared our serotypes to those from recently published studies in the UK^12–14^ and globally^15–19^. Overall, whilst there was some regional variation, findings supported the potential for O-antigen-targeted quadrivalent vaccines to contribute to substantial reductions in overall BSIs, as well as those caused by ESBL and MDR isolates, in both *E. coli* and *K. pneumoniae* (table 1). For *K. pneumoniae* they would likely provide a good level of protection against carbapenem-resistant and (genetically) hypervirulent isolates.

**Table 1.**
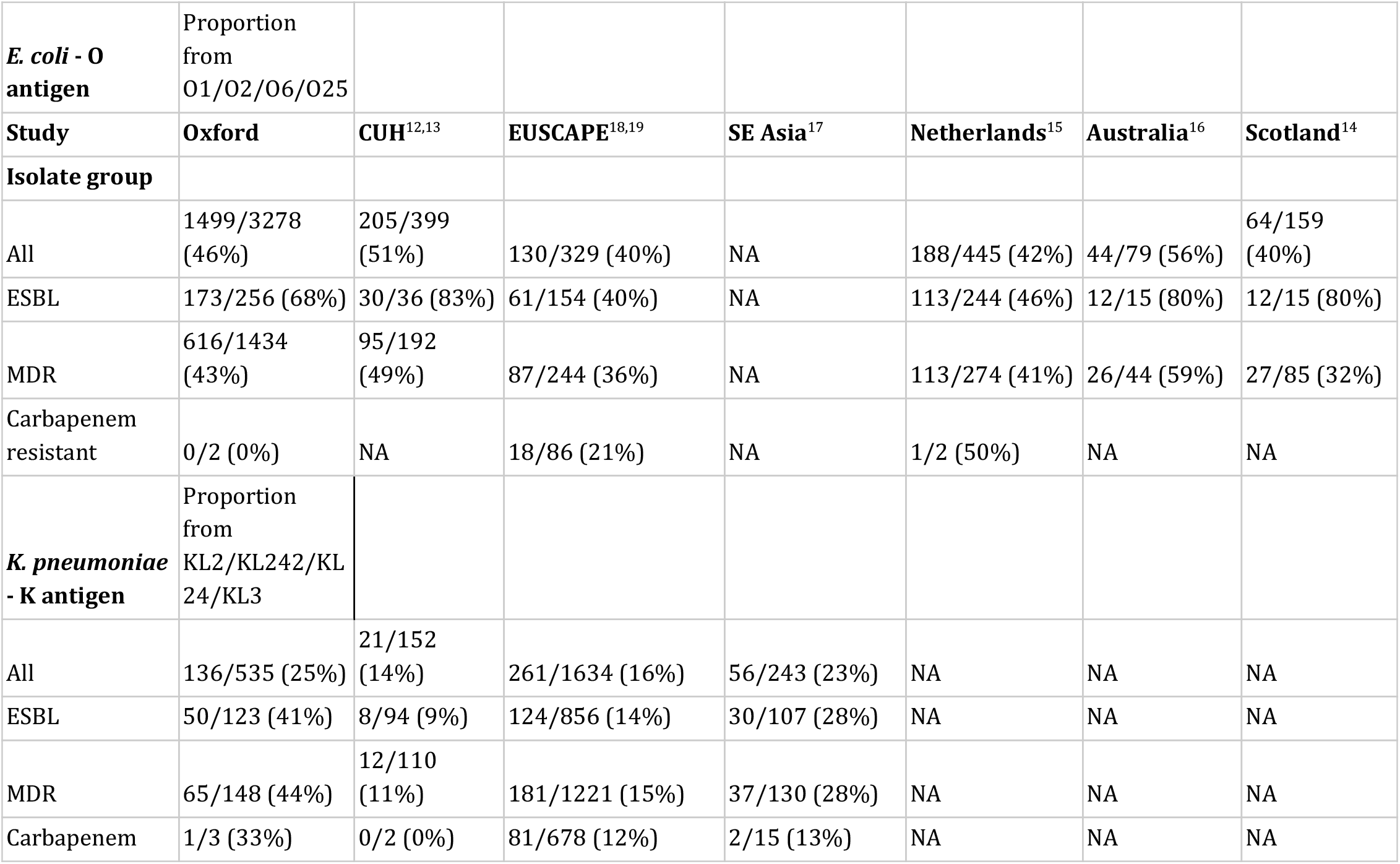
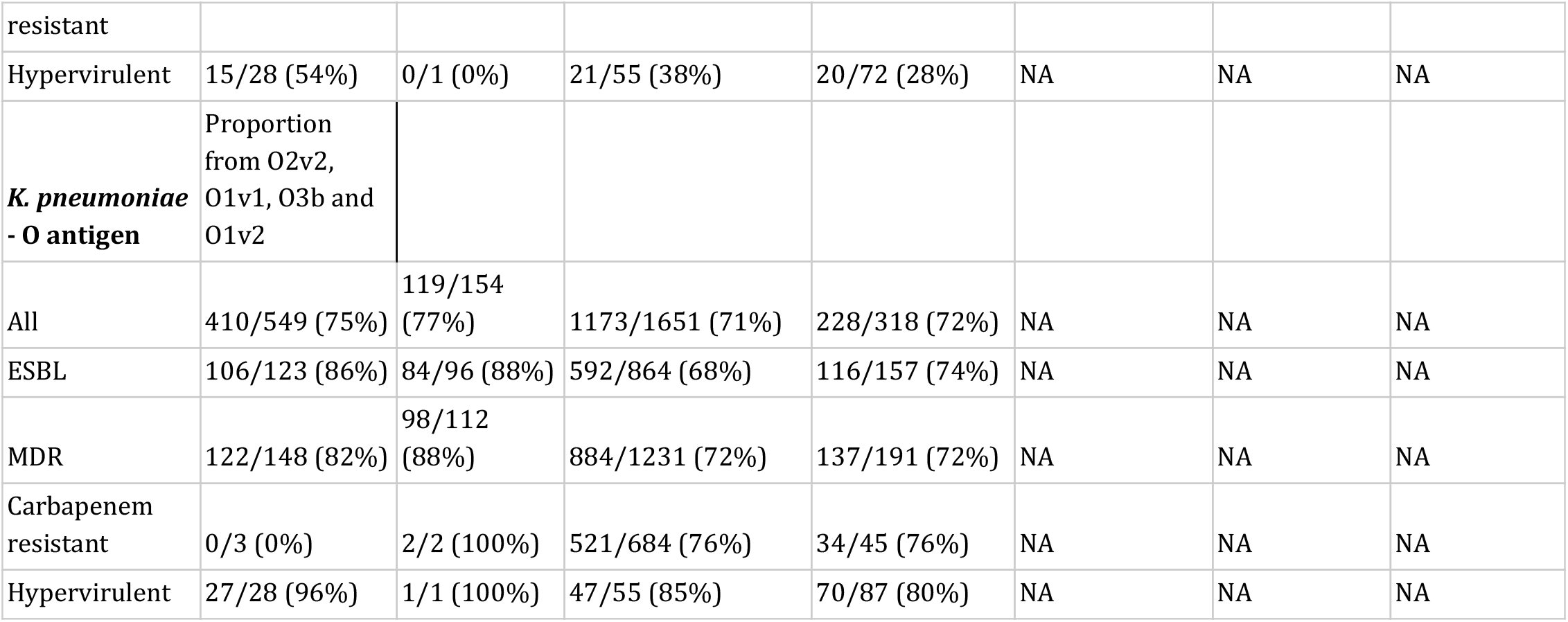
Comparison of the Oxford BSIs with those from similar national/regional studies. For each group we considered the proportion of isolates in each study carrying the four most prevalent antigens in the Oxford dataset to consider the hypothetical coverage a vaccine targeted at these might provide. CUH - Cambridge University Hospitals, EUSCAPE - The European Survey of Carbapenemase-Producing Enterobacteriaceae, SE Asia - South-east Asia. 3479 *E. coli* isolates from Oxford were sequenced of which 3278 passed Ectyper QC. 556 Oxford *K. pneumoniae* isolates were sequenced of which 535 (K-antigen)/549 (O-antigen) passed Kleborate QC. 1510 E. coli isolates from published datasets were included of which 1411 passed Ectyper QC. Of the 2168 *K. pneumoniae* isolates included from published datasets, 2029 (K-antigen)/2123 (O-antigen) passed Klerorate QC typing. The ESBL/MDR/Carbapenem resistant numbers are based on gene detection from Kleborate, including for Oxford isolates (in contrast to the main text where we report the results of *in vitro* drug susceptibility testing).

Conversely for *E. coli*, only 19/90 (21%) isolates carrying carbapenem resistance genes in all studies carried O-antigens covered by the ExPEC4V vaccine, with the O102 (n=18), O25 (n=11), O8 (n=11) and O160 (n=10) antigens predominating in these isolates. Reflecting this, only 13/90 isolates carrying carbapenem resistance genes belonged to the major group of sequence types (STs) (131/95/73/69) which are responsible for most (predominantly CA) *E. coli* BSIs reported previously^12,14,16^. This might indicate that these isolates do not originate from the community reservoir and may be associated with nosocomial transmission, as shown in *K. pneumoniae*^*19*^. Widespread dissemination of carbapenem resistance genes into STs associated with global epidemics of CA-MDR disease (eg ST131) would be a public health disaster and a vaccine may have a role in preventing this. In addition, preventing ESBL infections would reduce the carbapenem selection pressure.

Vaccines should also provide some protection against non-BSI infections which may evolve into BSIs, in particular urinary tract infections. As well as reducing morbidity and providing economic benefits, this could also be expected to lower antibiotic selection pressures against *E. coli* and *Klebsiella* spp.. The relatively much higher incidence of these primary infections vs BSIs may increase the economic viability of any prospective vaccine which, particularly for *Klebsiella spp*., may be a significant barrier to development given the difficulty in identifying groups of patients in which vaccination would be cost-effective^20^.

The main limitation of our study is that it is from a single region. To mitigate this we analysed several existing studies from the literature; however, in contrast to our long-term regional surveillance most of these were either deliberately enriched for MDR isolates or at high risk of bias due to unclear sampling methodologies. Whilst direct comparison with our data is therefore difficult, the overall findings appear to be reflected both nationally and globally.

Overall, a quadrivalent vaccine comprising the four most prevalent O-antigens for *E. coli* and *Klebsiella* spp. could theoretically have a significant impact on the incidence of BSIs and ESBL/MDR infections, as well as offering an alternative approach to help tackle the global epidemic of AMR in Enterobacteriaceae. Our findings support future development and subsequent efficacy studies for such vaccines, provided they are demonstrated to be safe and immunogenic. The substantial capsular antigen diversity observed for *Klebsiella* spp. BSIs would likely preclude this as a useful vaccine target. Surveillance following the implementation of any vaccine would be essential in identifying any evidence of strain replacement.

## Data Availability

All sequencing data is in the process of being uploaded to NCBI under project accession number PRJNA604975.

## Authors’ Notes

### Data availability

all sequencing data created for this study has been deposited in the NCBI under study accession number PRJNA604975.

## Acknowledgements

This work uses data provided by patients and collected by the UK’s National Health Service as part of their care and support. We thank all the people of Oxfordshire who contribute to the Infections in Oxfordshire Research Database.

Research Database Team: L Butcher, H Boseley, C Crichton, DW Crook, D Eyre, O Freeman, J Gearing (community), R Harrington, K Jeffery, M Landray, A Pal, TEA Peto, TP Quan, J Robinson (community), J Sellors, B Shine, AS Walker, D Waller. Patient and Public Panel: G Blower, C Mancey, P McLoughlin, B Nichols. We would like to thank the EUSCAPE consortium for making their data available in the public domain. We are also grateful for the efforts of the many biomedical and research scientists, in particular Dai Griffiths, who have assisted this project over the past 10 years.

